# ECG abnormalities are strongly associated with CVD outcomes in low-risk individuals using the PREVENT risk equation

**DOI:** 10.64898/2026.03.28.26349408

**Authors:** Mouhammad J Alawad, Elsayed Z. Soliman, Todd M. Brown, Philip Akinyelure oluakins, Hugo G. Quezada-Pinedo, Mohamed A. Mostafa, Mohan Satish, Parag Goyal, Orysya Soroka, Monika M. Safford

**Affiliations:** Weill Cornell Medicine, Department of Medicine, New York, NY; Wake Forest University School of Medicine, Department of Cardiovascular Medicine, Winston-Salem, NC; University of Alabama at Birmingham, Department of Medicine, Birmingham, AL; Department of Population Health Sciences, Duke University School of Medicine, Durham, NC; Epidemiological Cardiology Research Center (EPICARE), Department of Cardiovascular Medicine, Wake Forest University School of Medicine, Winston-Salem, NC; Weill Cornell Medicine, department of Cardiology, New York, NY

**Keywords:** ECG abnormalities, Minnesota Code, Cardiovascular disease, Coronary heart disease, Stroke, heart failure, primary prevention

## Abstract

**Background:** Resting electrocardiogram (ECG) is not currently recommended as part of cardiovascular disease (CVD) risk assessment, although accumulating evidence suggests a potential role.

**Objective:** To examine the association between ECG abnormalities and incident CVD events as assessed by the 2023 Predicting Risk of Cardiovascular Disease Events (PREVENT) equations.

**Design:** Secondary data analysis from the REasons for Geographic And Racial Differences in Stroke (REGARDS) prospective cohort, including study participants without a baseline CVD.

**Exposure:** ECG abnormalities were classified by Minnesota Code (MC) as normal, any minor, or major abnormality at baseline (2003-2007).

**Outcome:** Participants were followed for expert adjudicated incident CVD events through December 31, 2021.

**Results:** Among 19,173 participants (mean age at baseline of 63.7 years; 57.8% were female). According to the PREVENT risk equations, 39.4% were classified as <7.5% 10-year risk CVD risk, 44.6% as 7.5-20% risk, and 16.0% as >20% risk. Overall, 47.0% had normal ECG, 44.0% had any minor abnormality, and 9.0% had any major abnormality. During follow-up, CVD events occurred in 12.4% of participants with normal ECG, 17.0% of those with any minor abnormality, and 25.4% of those with any major abnormality. Compared to those without ECG abnormality, the adjusted HR for incident CVD were 1.19 (95% CI 1.10-1.29) for any minor abnormality, and 1.53 (1.36-1.72) for any major ECG abnormality. In the <7.5% risk group, 43.6% had at least one ECG abnormality; in this risk group compared to those without ECG abnormality, the HR for incident CVD associated with any major ECG abnormality, present in 5.0% of the <7.5% risk group, was 1.87 (95% CI 1.34-2.62), The HR for any minor ECG abnormalities, present in 38.6% was 1.13 ( 95% CI 0.93 - 1.37).

**Conclusion:** ECG abnormalities were associated with risk of CVD events across PREVENT risk groups. A substantial proportion of low-risk participants (according to the PREVENT equation) had ECG abnormalities and associated elevated risk. This supports the potential for using ECG to identify a subgroup of low-risk patients who may benefit from more aggressive primary prevention especially with major ECG abnormalities. Addition of electrocardiographic evaluation to the PREVENT risk equations may improves cardiovascular risk discrimination.

## INTRODUCTION

Cardiovascular diseases (CVD) are the main cause of death in the United States (US) and globally,^1,2^ and after a long decline over the past 30 years, deaths attributable to CVD started to rise again in the last decade.^2,3^ CVD burden is huge, it affects quality of life, morbidity, and healthcare expenditure.^3^

As the US population ages, CVD preventive strategies become increasingly essential. Multiple risk factors have been identified and risk factors modification has been targeted to lower CVD risk.^4,5^ Various equations and risk scores have been developed to predict CVD risk, but each has its own limitation based on populations and specific circumstances for individuals.^5,6^

Unfortunately even those classified as low risk, can develop CVD events; in fact, 15% of patients with acute myocardial infarction (MI) have no traditional risk factors^7^. This supports the need for to identify subgroups of patients at low risk for future CVD events, who could potentially benefit from a targeted intervention.

Resting electrocardiogram (ECG) is a simple, non-invasive, and widely available test,^8^ but it is not currently part of CVD risk stratification.^9^ The United States Preventive Services Task Force (USPSTF) recommended in 2018 against screening with resting or exercise ECG to prevent CVD events in asymptomatic adults at low risk of CVD events (grade D evidence), but stated that the current evidence is insufficient to assess the balance of benefits and harms of screening in asymptomatic adults at intermediate or high risk of CVD events.^10^ Nevertheless, ECG abnormalities may have a potential for screening and risk stratification in otherwise healthy individuals ^11,12^. In fact, since the 2018 recommendation, emerging evidence suggests that ECG screening may in fact hold a promise. A 2024 study by Yagi et al showed that ECG abnormalities were associated with CVD outcomes in 3.5 million working-age adults in Japan, regardless of baseline CVD risk^13^. The main outcome was a combination of all-cause mortality or hospitalization for CVD (stroke, heart failure, or acute MI). However, it is unclear to what extent the study’s findings apply to other countries. A US-based study using the Third National Health and Nutrition Examination Survey in adults free of CVD and major ECG abnormalities at baseline showed that as the number of minor ECG abnormalities increased, the risk of CVD mortality increased, with a 13% increase in risk with each additional minor ECG abnormality.^14^ This study was based on data collected between 1988 to 1994, and it examined only CVD mortality.^14^

In this study, we used data from the national REasons for Geographic And Racial Difference in Stroke (REGARDS) cohort to examine the relationship between baseline ECG findings among individuals free of CVD at baseline in associations with future incident CVD events and CVD mortality while using the PREVENT equation for risk stratification. We notably looked at <7.5% risk group. REGARDS cohort is contemporary, with subjects recruited between 2003-7, has rigorously adjudicated CVD endpoints and causes of death, and has national reach. We hypothesized that ECG abnormalities are associated with risk of CVD events, in all CVD risk groups as assessed by the 2023 Predicting Risk of Cardiovascular Disease Events (PREVENT) equations.

## METHODS

### Study Population

The REGARDs study is a prospective national biracial cohort designed to define regional and racial differences in stroke mortality. Details are provided elsewhere,^15^ but in brief, from 2003 to 2007, 30239 community-dwelling individuals aged 45 years and older were enrolled using commercially available lists. The sampling design over recruited from the Southeastern Stroke Belt states, with 50% residing in Louisiana, Mississippi, Alabama, Georgia, North and South Carolina, Tennessee, and Arkansas, and 50% residing in the other contiguous US states. Within the Stroke Belt, one third of the sample resided in the Stroke Buckle, consisting of the coastal areas of North and South Carolina and Georgia. Self-reported race other than Black or white, inability to speak English, active cancer treatment, and inability to complete a survey and in-home exam were exclusions. Baseline data included a medical history and in-home exam collecting anthropometrics, blood and urine samples, and an ECG. Participants are contacted by telephone at 6-months intervals to detect potential CVD events with subsequent retrieval of medical records for adjudication. Follow-up is ongoing. Written informed consent was obtained from all subjects, and the institutional review boards at all participating institutions approved the protocol.

For this study, participants free of CVD at baseline were included. CVD was ascertained through self-reported physician-diagnosed stroke, heart attack, coronary revascularization, or carotid revascularization.^15^ Participants with evidence of an old MI on the baseline ECG were also excluded. We used an algorithm developed by Goyal et al to define a HF-free phenotype (negative predictive value 99%).^16^

### Main exposure: ECG abnormalities

Electrocardiograms obtained during the in-home baseline visit were transmitted for centralized interpretation at the Epidemiological Cardiology Research Center (Wake Forest School of Medicine, Winston-Salem, NC, USA). All tracings were read by trained analysts blinded to clinical data and study outcomes. ECG findings were classified according to the Minnesota Code (MC).^17^ Participants with only minor abnormalities were categorized as having a minor ECG abnormality, whereas those with any major abnormality—regardless of the presence of coexisting minor abnormalities—were categorized as having a major ECG abnormality. Major ECG abnormalities included: Major ventricular conduction defect; definite myocardial infarction (defined as the presence of major Q wave abnormalities); possible myocardial infarction (defined as the presence of minor Q-QS wave plus major ST-T abnormalities); major isolated ST-T abnormalities; left ventricular hypertrophy plus major ST-T abnormalities; major atrio-ventricular conduction abnormalities; major QT prolongation (QTI≥116% or JTI if QRS ≥120 milliseconds [ms]), pacemaker, and other major arrhythmias. Minor ECG abnormalities included: Minor isolated Q-QS waves; minor isolated ST-T abnormalities; high R waves; ST segment elevation; incomplete right bundle branch block; minor QT prolongation (QTI≥112% or JTI if QRS≥120 ms); short PR interval; left axis deviation; right axis deviation; frequent ventricular premature beats; and other minor abnormalities (see supplemental Table 5).

### Outcomes

The outcome of this study was incident CVD, defined as incident CHD (fatal and nonfatal MI), heart failure, stroke, or incident CVD death (primary underlying cause of death CHD, stroke, or HF).

Nonfatal MI was adjudicated based on a clinical presentation consistent with ischemia, biomarkers, and/or ECG findings consistent with of ischemia. Biomarker patterns included a rising and/or falling patterns of cardiac troponin or creatine kinase-MB with a peak more than two times the upper limit of normal over a period of at least 6 hours.^18^

Fatal MI was defined as death within 28 days of hospital admission for definite or probable MI or death within 6 hours of hospital admission with symptoms/signs with absent or nondiagnostic ECG or biomarkers, or evidence indicating likely sudden death.^19^ Adjudication of cause of death used all available information including medical history, hospital records, autopsy reports, death certificates, interviews with next of kin or proxies, and the National Death Index.

Heart failure was adjudicated based on a hospitalized clinical presentation consistent with HF, treatment consistent with HF, b-type natriuretic protein elevation, and imaging findings.^20^

Stroke was adjudicated using the World Health Organization definition of the acute onset of a focal neurological defect consistent with ischemia lasting more than 24 hours or resulting in death before 24 hours; clinical strokes were adjudicated if symptoms lasted less than 24 hours but imaging findings on magnetic resonance imaging were consistent with an acute stroke.

Transient ischemic attacks (TIA) were not included as stroke events.^21^

### Covariates

Covariates were selected based on confounding and to account for study sampling and included sociodemographic factors, health behaviors, chronic medical conditions, inflammatory markers and medications. These were: age, sex, REGARDS region, annual household income, education, hypertension (HTN), treatment for HTN, diabetes, insulin treatment, high sensitive C reactive protein (hsCRP), albumin-to-creatinine ratio, eGFR, total cholesterol, HDL cholesterol, cigarette smoking, lack of physical activity, BMI, adherence to the Mediterranean diet, depressive symptoms, and family history of premature CVD). Only variables not present in the PREVENT equations were entered into multivariable models (see below).

### Statistical Analysis

We categorized the sample into the 3 ECG groups defined above (normal, any minor, and major abnormalities). We used the PREVENT base equation for CVD outcomes to classify participants as low risk (<7.5%) intermediate risk (7.5-20%), and high risk (>20%) of experiencing a CVD event in the next 10 years.^22^ The PREVENT base equations included age, sex, cigarette smoking, total cholesterol, HDL cholesterol, systolic blood pressure, blood pressure medication treatment, diabetes, eGFR, using statins, body mass index.^22^ We then compared the characteristics of the 3 ECG groups on sociodemographic factors, CVD risk factors, and CVD risk groups. We reported the proportion of each CVD risk group that experienced any CVD event.

Next, we constructed Kaplan-Meier (KM) survival curves for CVD events classifying individuals into the 3 ECG groups. We used the log-rank test to determine the statistical significance of differences in the curves. We created KM curves for the whole sample as well as for each CVD risk group separately.

We then examined the association between the baseline ECG category and future CVD events. We used an analogous approach for each analysis. We used Cox proportional hazards regression to examine the risk of relevant outcome associated with ECG category. With participants censored at the date of an outcome, date of withdrawal from the study, date of death, or December 31, 2021, whichever came first. We first constructed Cox proportional hazard models to examine the age and sex adjusted model for the association between the relevant outcome and each of the non-normal groups of ECG categories compared to no ECG abnormalities, generating hazard ratios (HR) and 95% confidence intervals (CI). We then added sociodemographic, health behaviors, chronic medical conditions, inflammatory markers and medications variables to the model and observed changes in the magnitude and directionality of the parameter estimate for the main exposure variable (ECG risk group variable). We did not adjust for covariates that are identical in the PREVENT base equation to avoid double adjusting for the same components. We accounted for the competing risk of non-CVD death using the method described by Fine and Gray.^23^ For all analyses described above, the proportional hazards assumption was examined using Schoenfeld residuals.

To examine differences by sex, age (<65 and ≥65 years), and CHD/CVD risk classification group, we tested interaction terms and stratified models if the p-value for the interaction was <0.10.

Because of the interest in whether routine ECG could be used as an addition to current risk stratification approaches for low-risk groups, we stratified the model on CVD risk groups regardless of interaction term p-value. Due to concerns about underutilization of health services by socially marginalized and racialized groups, we conducted a separate analysis including race in the model, and examined interactions by race, again stratified if the p-value for the interaction was <0.10.

We implemented multiple imputation by chained equations to minimize bias and maximize sample size. All tests were 2-tailed and p<0.05 was used as the threshold for statistical significance except in the test for interactions, where the p-value was set at <0.10. Analyses were conducted using SAS version 9.4 (SAS Institute, Cary, NC) and Stata, version 18.0 (StataCorp, College Station, TX).

## RESULTS

### Sample characteristics

Out of 30,239 REGARDS participants, 21,131 participants had no evidence of CVD at baseline with available ECG information. The final cohort included 19,173 participants with all information needed to calculate the CVD risk by PREVENT base equation. Median follow-up time was 13.3 years (IQR 7.0-16.1). The mean age at baseline was 63.7 years and 57.8% were female, 40.2% self-identified as having Black race, 10.1% reported less than a High School education, and 43.8% reported annual household income <$35,000 (Table 1 and full details in Supplementary Table 1). The prevalence of HTN was 70.4%, 53.6% were receiving anti-HTN medications, 19.9% had diabetes, and 24.3% were on statins. In the study sample, 9,060 (47%) individuals had a normal ECG, 8,384 (44%) had any minor abnormality, and 1.729 (9%) had a major ECG abnormality.

**Table 1:**
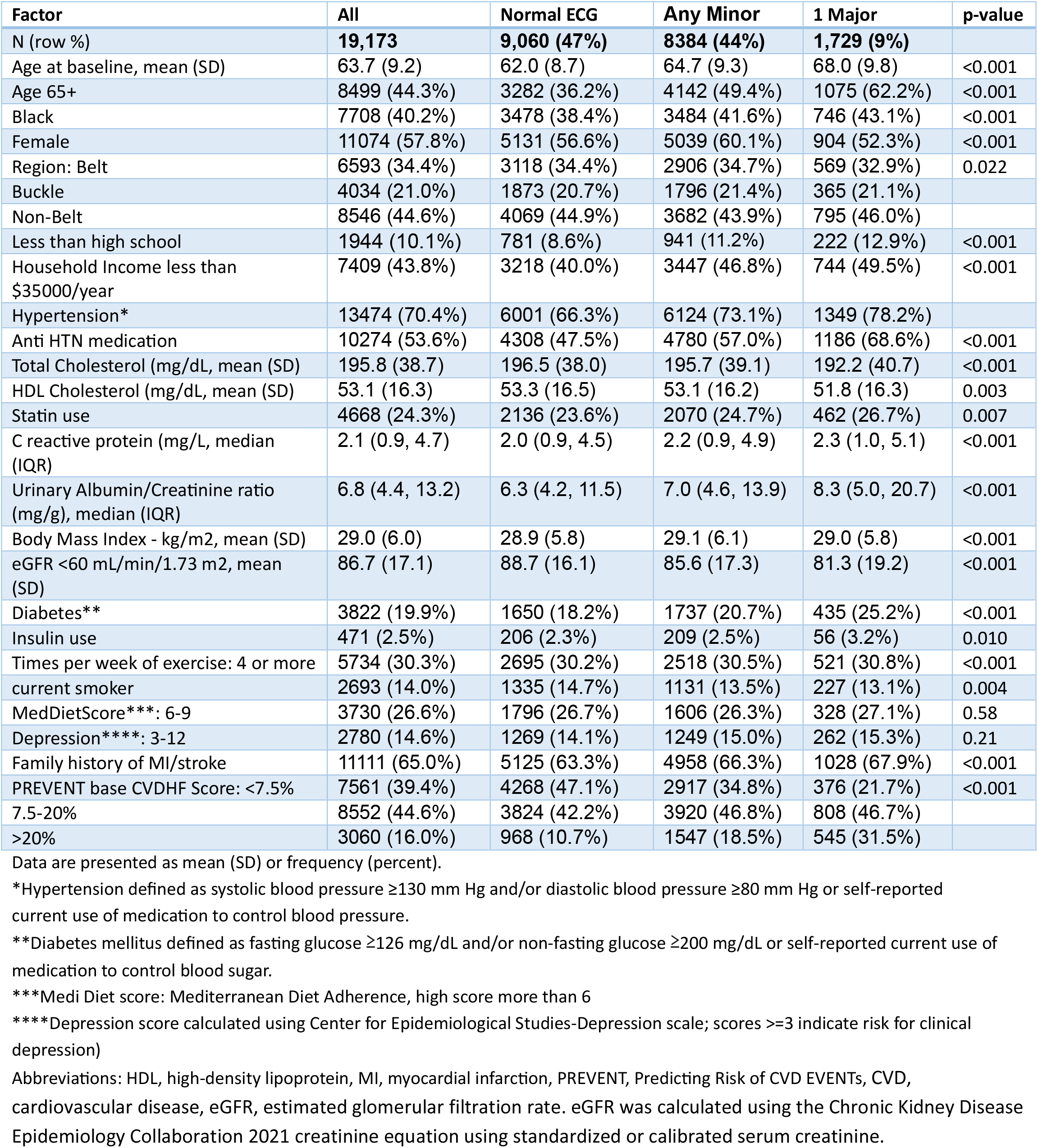
Baseline Characteristics of REGARDS Study Participants stratified by ECG categories.

| Factor | All | Normal ECG | Any Minor | 1 Major | p-value |
| --- | --- | --- | --- | --- | --- |
| N (row %) | **19,173** | **9,060 (47%)** | **8384 (44%)** | **1,729 (9%)** |  |
| Age at baseline, mean (SD) | 63.7 (9.2) | 62.0 (8.7) | 64.7 (9.3) | 68.0 (9.8) | <0.001 |
| Age 65+ | 8499 (44.3%) | 3282 (36.2%) | 4142 (49.4%) | 1075 (62.2%) | <0.001 |
| Black | 7708 (40.2%) | 3478 (38.4%) | 3484 (41.6%) | 746 (43.1%) | <0.001 |
| Female | 11074 (57.8%) | 5131 (56.6%) | 5039 (60.1%) | 904 (52.3%) | <0.001 |
| Region: Belt | 6593 (34.4%) | 3118 (34.4%) | 2906 (34.7%) | 569 (32.9%) | 0.022 |
| Buckle | 4034 (21.0%) | 1873 (20.7%) | 1796 (21.4%) | 365 (21.1%) |  |
| Non-Belt | 8546 (44.6%) | 4069 (44.9%) | 3682 (43.9%) | 795 (46.0%) |  |
| Less than high school | 1944 (10.1%) | 781 (8.6%) | 941 (11.2%) | 222 (12.9%) | <0.001 |
| Household Income less than \$35000/year | 7409 (43.8%) | 3218 (40.0%) | 3447 (46.8%) | 744 (49.5%) | <0.001 |
| Hypertension* | 13474 (70.4%) | 6001 (66.3%) | 6124 (73.1%) | 1349 (78.2%) |  |
| Anti HTN medication | 10274 (53.6%) | 4308 (47.5%) | 4780 (57.0%) | 1186 (68.6%) | <0.001 |
| Total Cholesterol (mg/dL, mean (SD) | 195.8 (38.7) | 196.5 (38.0) | 195.7 (39.1) | 192.2 (40.7) | <0.001 |
| HDL Cholesterol (mg/dL, mean (SD) | 53.1 (16.3) | 53.3 (16.5) | 53.1 (16.2) | 51.8 (16.3) | 0.003 |
| Statin use | 4668 (24.3%) | 2136 (23.6%) | 2070 (24.7%) | 462 (26.7%) | 0.007 |
| C reactive protein (mg/L, median (IQR) | 2.1 (0.9, 4.7) | 2.0 (0.9, 4.5) | 2.2 (0.9, 4.9) | 2.3 (1.0, 5.1) | <0.001 |
| Urinary Albumin/Creatinine ratio (mg/g), median (IQR) | 6.8 (4.4, 13.2) | 6.3 (4.2, 11.5) | 7.0 (4.6, 13.9) | 8.3 (5.0, 20.7) | <0.001 |
| Body Mass Index - kg/m2, mean (SD) | 29.0 (6.0) | 28.9 (5.8) | 29.1 (6.1) | 29.0 (5.8) | <0.001 |
| eGFR <60 mL/min/1.73 m2, mean (SD) | 86.7 (17.1) | 88.7 (16.1) | 85.6 (17.3) | 81.3 (19.2) | <0.001 |
| Diabetes** | 3822 (19.9%) | 1650 (18.2%) | 1737 (20.7%) | 435 (25.2%) | <0.001 |
| Insulin use | 471 (2.5%) | 206 (2.3%) | 209 (2.5%) | 56 (3.2%) | 0.010 |
| Times per week of exercise: 4 or more | 5734 (30.3%) | 2695 (30.2%) | 2518 (30.5%) | 521 (30.8%) | <0.001 |
| current smoker | 2693 (14.0%) | 1335 (14.7%) | 1131 (13.5%) | 227 (13.1%) | 0.004 |
| MedDietScore***: 6-9 | 3730 (26.6%) | 1796 (26.7%) | 1606 (26.3%) | 328 (27.1%) | 0.58 |
| Depression****: 3-12 | 2780 (14.6%) | 1269 (14.1%) | 1249 (15.0%) | 262 (15.3%) | 0.21 |
| Family history of MI/stroke | 11111 (65.0%) | 5125 (63.3%) | 4958 (66.3%) | 1028 (67.9%) | <0.001 |
| PREVENT base CVDHF Score: <7.5% | 7561 (39.4%) | 4268 (47.1%) | 2917 (34.8%) | 376 (21.7%) | <0.001 |
| 7.5-20% | 8552 (44.6%) | 3824 (42.2%) | 3920 (46.8%) | 808 (46.7%) |  |
| >20% | 3060 (16.0%) | 968 (10.7%) | 1547 (18.5%) | 545 (31.5%) |  |

Data are presented as mean (SD) or frequency (percent).
\*Hypertension defined as systolic blood pressure $\geq 130$ mm Hg and/or diastolic blood pressure $\geq 80$ mm Hg or self-reported current use of medication to control blood pressure.
\*\*Diabetes mellitus defined as fasting glucose $\geq 126$ mg/dL and/or non-fasting glucose $\geq 200$ mg/dL or self-reported current use of medication to control blood sugar.
\*\*\*Medi Diet score: Mediterranean Diet Adherence, high score more than 6
\*\*\*\*Depression score calculated using Center for Epidemiological Studies-Depression scale; scores $\geq 3$ indicate risk for clinical depression)
Abbreviations: HDL, high-density lipoprotein, MI, myocardial infarction, PREVENT, Predicting Risk of CVD EVENTS, CVD, cardiovascular disease, eGFR, estimated glomerular filtration rate. eGFR was calculated using the Chronic Kidney Disease Epidemiology Collaboration 2021 creatinine equation using standardized or calibrated serum creatinine.

### ECG categories by strati of risk classification

Table 2 shows the distribution of participants in the 3 PREVENT score categories within each ECG category. Using 7.5% 10-year risk to define low risk, 39.4% of the sample was classified as low risk (i.e. <7.5%), 44.6% as intermediate risk (7.5-20%), and 16% as high risk (>20%). Among those at low risk, 43.6% had at least one ECG abnormality, 38.6% had any minor ECG abnormality, and 5.0% had any major ECG abnormality. Supplementary Table 2 further classifies the sample based on presence of only one minor ECG abnormality, one or more minor abnormality, and any major abnormality, demonstrating that 20.6% of the sample had a single minor ECG abnormality.

**Table 2:** ECG categories in each PREVENT risk score classification.

|  | All | ECG classification |  |  |  |  |  |
| --- | --- | --- | --- | --- | --- | --- | --- |
|  |  | Normal |  | Minor |  | Major |  |
|  | N | N | row % | N | row % | N | row % |
| **All** | 19173 | 9060 | 47 | 8384 | 44 | 1729 | 9 |
| **PREVENT Base CVD Score:** |  |  |  |  |  |  |  |
| **Low risk (<7.5%)** | 7561 | 4268 | 56.4 | 2917 | 38.6 | 376 | 5 |
| **intermediate risk (7.5-20%)** | 8552 | 3824 | 44.7 | 3920 | 45.8 | 808 | 9.5 |
| **High risk (>20%)** | 3060 | 968 | 31.6 | 1547 | 50.6 | 545 | 17.8 |

### Kaplan-Meier survival curves

During follow-up, 15.6% of the sample had a CVD event (Figure 1). CVD events occurred in 12.4% of the sample with normal ECG, 17% of those with any minor abnormality group, and 25.4 % of the major ECG abnormality group. CVD events occurred in 6.4% in the <7.5% low-risk group, 18.7% of those in the 7.5-20% risk group, and 29.6% of those in the >20% risk group (Figure 2). Survival curves for 1 minor, 2 or more minor, and any major ECG abnormalities are shown in supplemental Figure 1, demonstrating similar risk for those with 1 minor or 2 or more minor abnormalities. Supplemental Figure 2 shows survival curves for the three risk strata with the 4 ECG categories (normal, 1 minor, 2 or more minor, any major), demonstrating that in the lower risk groups the 1 minor and 2 or minor categories had similar survival. Survival curves stratified on sex and age are shown in supplemental figure 3 and 4, respectively, showing similar patterns in these groups as in the overall sample. Of the 3261 CVD events, 1584 (48.6%) were CHD events, 1184 (36.3%) were stroke events, and 493 (15.1%) were HF events.

**Figure 1:**
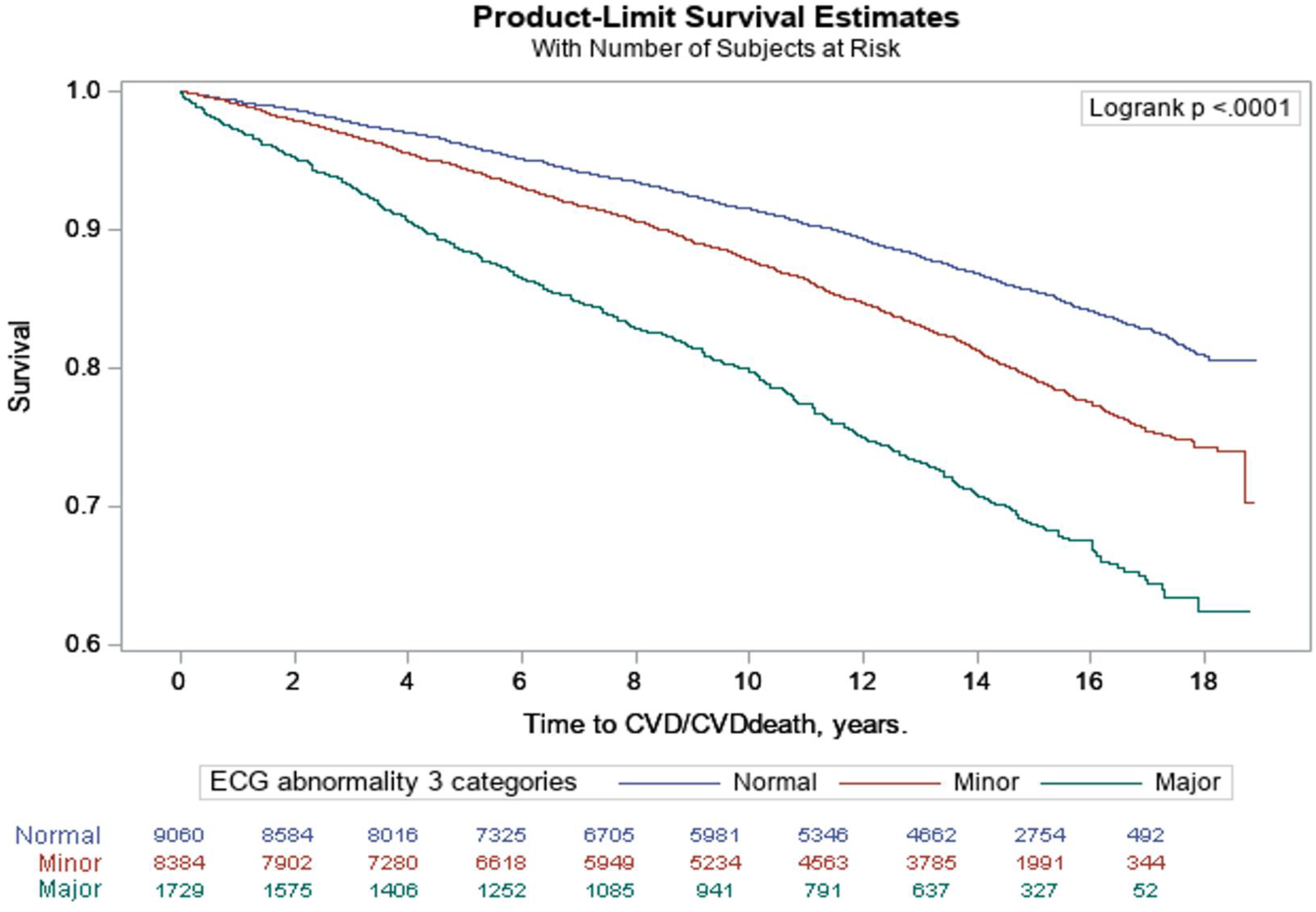
Kaplan-Meier Survival Curves for CVD/CVD death incidents during follow-up (2003-2021) by ECG categories in the overall population.

**Figure 2:**
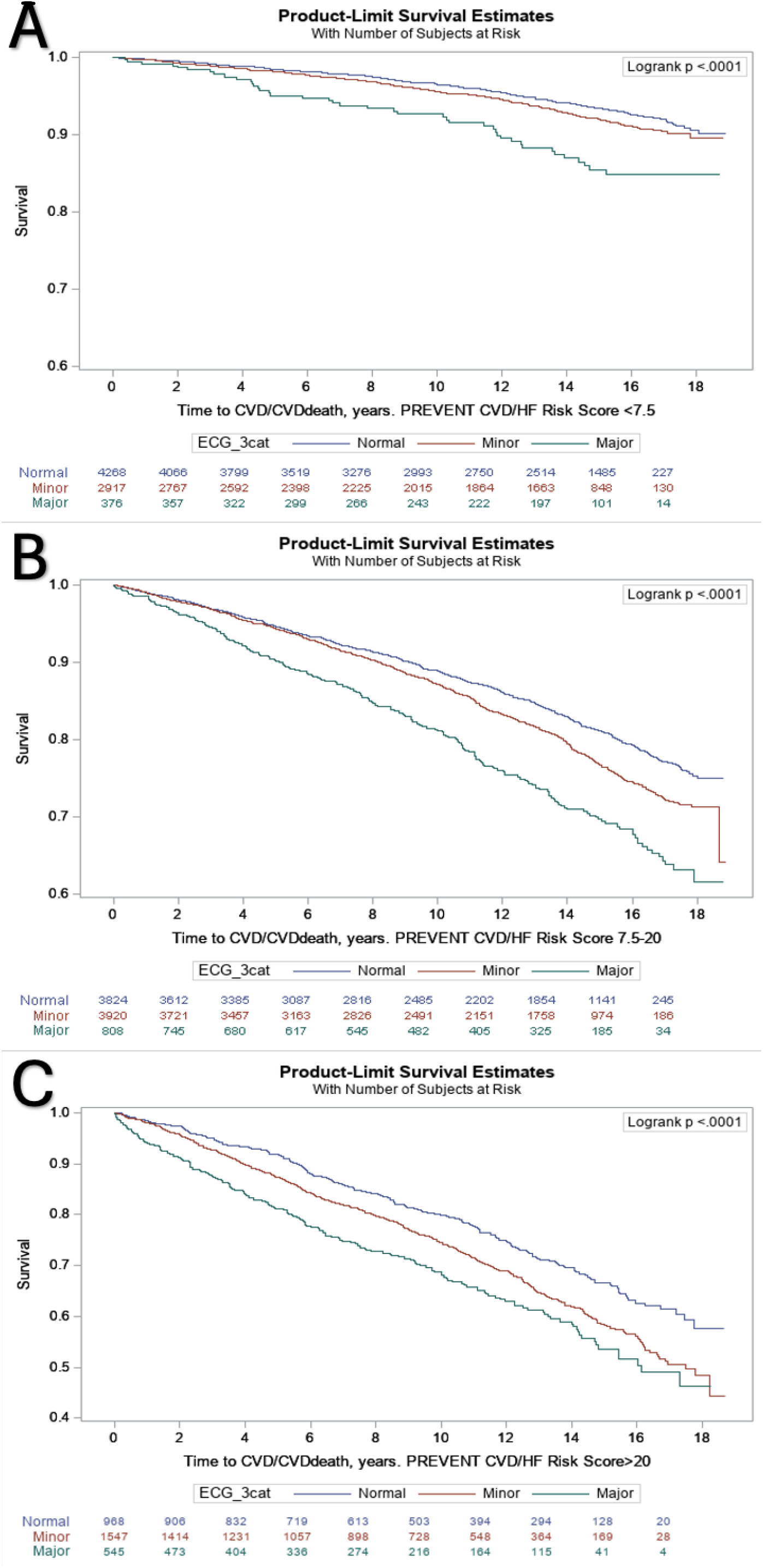
Kaplan-Meier probability of being CVD/CVD death free during follow-up (2003-2021) by baseline electrocardiogram malities for 3 categories of PREVENT score: A - <7.5%, B – 7.5-20%, C - >20%.

### Multivariable modeling results

The Cox proportional hazard models showed that compared to those with normal ECGs, individuals with any ECG abnormality at baseline had higher risk of CVD events in both age-sex adjusted and fully adjusted models (Table 3). The age-sex adjusted models HR for any minor ECG abnormality was 1.26 (95% CI 1.16-1.36), and for any major ECG abnormality was 1.68 (95% CI 1.49-1.88). The relationship remained significant after adjusting for covariates: the HR for any minor ECG abnormality was 1.19 (95% CI 1.10-1.29), and for any major ECG abnormality was 1.53 (95% CI 1.36-1.72).

**Table 3:** CVD outcome group: COX proportional hazard ratios (95% CI) for incident CVD by ECG categories as exposure ared to normal ECGs, age, sex adjusted model in the upper followed by fully adjusted models followed by fully ed imputed model in the lower panel, stratified according to PREVENT score of <7.5%, 7.5 – 20% and >20%.

| ECG categories | overall | PREVENT risk <7.5%<br>(Low risk) | PREVENT risk 7.5-20%<br>(Intermediate risk) | PREVENT risk >20%<br>(High risk) |
| --- | --- | --- | --- | --- |
| **Age/sex adjusted model** |  |  |  |  |
| **Any Minor** | **1.26 (1.16-1.36)** | 1.19 (0.98-1.44) | **1.19 (1.07-1.32)** | **1.25 (1.07-1.45)** |
| **Any Major** | **1.68 (1.49-1.88)** | **2.08 (1.49-2.89)** | **1.65 (1.41-1.93)** | **1.45 (1.2-1.77)** |
| **Normal** | ref | ref | ref | ref |
| **Fully adjusted models\*** |  |  |  |  |
| **Any Minor** | **1.19 (1.10 - 1.29)** | 1.13 (0.93 - 1.37) | **1.15 (1.03 - 1.28)** | **1.22 (1.05 - 1.43)** |
| **Any Major** | **1.53 (1.36 - 1.72)** | **1.87 (1.34 - 2.62)** | **1.53 (1.30 - 1.79)** | **1.40 (1.15 - 1.70)** |
| **Normal** | ref | ref | ref | ref |

\*Adjusted for age, sex, REGARDS region, annual household income, education, insulin treatment, CRP, albumin-to-creatinine ratio, physical activity, BMI, adherence to the Mediterranean diet, depressive symptoms, and family history of premature CVD.

Table 3 also shows HR for models stratified on PREVENT score (low risk<7.5%, intermediate 7.5-20%, high risk>20%). We found that individuals with low risk had a stronger association with any major ECG abnormality compared to other risk groups: the HR for any major ECG abnormality in the low-risk group was 2.08 (95% CI 1.49-2.89) and remained strong after adjusting for covariates (HR 1.87, 95% CI 1.34-2.62). The odds for any minor ECG abnormality in the low-risk group were of a similar magnitude as for the intermediate risk group but not statistically significant . The HR for 1 minor ECG abnormality, 2 or more minor abnormalities, vs. no abnormality are available in supplementary table 3, the HR for the age-sex adjusted model for 1 minor abnormality was 1.31 (95% CI 1.04-1.65) which became nonsignificant with full adjustment (HR 1.20, 95% CI 0.95-1.53) (supplementary Table 3). Of note, the HR associated with 1 minor abnormality is bigger in the lowest risk group than in the other risk groups and bigger than HR of ≥ minor abnormalities (supplementary Table 3).

The p-value for interactions with age (p=0.02) and sex (p=0.02) supported stratified analyses, which revealed that findings were largely similar for older and younger individuals, and for men and women (supplementary tables 4, supplementary figures 3 and 4). no effect modification observed for race groups.

## DISCUSSION

In this large, diverse, national, contemporary sample of community-dwelling adults with 13 years median follow-up, we observed 1) a significant association between baseline ECG abnormalities and CVD events. 2) ECG abnormalities are present in > 43% of people with PREVENT score of <7.5%. 3) Notably, the HR for CVD events with ECG abnormalities was greater in the low-risk group compared to the intermediate and higher risk groups, suggesting that ECG may be useful in further risk stratifying this large group. Our findings contribute to the growing evidence that ECG abnormalities may be particular useful providing incremental value specifically among adults considered to be at lower risk for CVD events.^13,24,25^

As CVD burden is increasing among younger populations, optimizing risk stratification in apparently low-risk groups (which is primarily compromised of younger adults) is becoming increasingly important.^24^ A study from Korea examined minor ECG abnormalities score and coronary arteries calcification (CAC) score and found a significant association between ECG abnormalities and a higher prevalence of subclinical CAC in a relatively young and low-risk population.^24^ Another study from China that examined ECG abnormalities and CVD events showed that 57.5% of people at low risk for CVD (<5% according to Chinese guidelines) had ECG abnormalities,^25^ comparable to our findings of 43.6% in the <7.5% low-risk group using PREVENT.

Resting ECG is not currently recommended for CVD risk stratification in the general population.^26^ Over the past 2 decades, accumulating evidence suggests that minor and major ECG abnormalities are independently associated with incident CHD and CVD^27,28^, cardiovascular mortality, and all-cause mortality,^29^ and in subgroups defined by sex, age, and race.^27,29–31^ Similar to prior studies,^30^ we show here that ECG abnormalities are common; present in over half of this sample of people without clinically evident CVD. Others have similarly described non-specific ECG abnormalities in the general population^31,32^ without clinically detected CVD.^33^ Although non-specific, such ECG changes may represent a marker for subclinical and/or prior myocardial injury, which elevates risk.^14,30^ In the literature certain ECG abnormalities are more strongly associated with CVD outcomes than others.^32,34^ For example, isolated nonspecific ST-segment, T-wave abnormalities (NSTTA) are significantly associated with increased risk of subclinical atherosclerosis and coronary death.^31,35^ Repolarization abnormalities have been found to be independent predictors for CHD death and sudden cardiac death.^34^

According to the 2019 ACC/AHA Guideline on the Primary Prevention of Cardiovascular Disease, in adults with <7.5% 10-year ASCVD risk or intermediate (≥7.5% to <20%) 10-year ASCVD risk, it is reasonable to use additional risk-enhancing factors to guide decisions about preventive interventions. Based on accumulating evidence from our study and others,^24,25^ ECG abnormalities may be additional strategy to guide these discussions in low risk profile individuals.

Studies that employed ECG abnormalities in risk stratification are being reported,^36^ with some authors having used ECG abnormalities to re stratify CVD risk profiles^9^ and others have examined whether they can improve the performance of risk scores like the Framingham and the European SCORE risk score.^30,37^ Kim, et al, showed that combined ECG abnormalities were better predictors of CVD death than conventional CVD risk factors and the 10-year ASCVD risk equation.^38^ The Minnesota Code is a valuable resource for classifying ECG abnormalities, with a possible role on prognostication and improving risk prediction.^39^ We used the Minnesota Code and our findings are consistent with previous studies.^13,14,40^ Because clinical applications of the MC may be challenging due to its details and complexity, artificial intelligence could be useful in this regard; recent studies showed utility to detect atrial fibrillation^41^ and systolic ventricular dysfunction^42^ with deep learning algorithms. Using PREVENT risk equations with ECG abnormalities may offer an opportunity to identify a group of individuals who may benefit from preventive measures.

Strengths of our study include its large, national, biracial sample, long follow-up period, expert adjunction of events following national guidelines, rigorously collected data, and the high likelihood of generalizability of findings given the national scope. Because we modeled our study design on Yagi et al, we were able to validate their results in a US sample. However, the results of our study need to be interpreted cautiously in the context of several limitations. The observational nature of the study warrants caution in drawing causal inferences. Although the REGARDS cohort included a large sample of Black participants, it did not include Latinos and other groups living in the US, warranting studies that do include these populations. As in any research study, participants may not be representative of the general population. Not all participants had ECG available. Some participants had missing data, which we compensated for by using multiple imputation. Some covariates were self-reported which may have introduced bias.

In conclusion, using the Minnesota code to classify ECG abnormalities, we found an association between ECG abnormalities and incident CVD events across all PREVENT risk groups, including the lowest risk group. This association was most striking in the lowest-risk group with major ECG abnormalities, adding to accumulating evidence that supports a potential role of ECG abnormalities in CVD risk stratification.

## Data Availability

All data produced in the present study are available upon reasonable request to the authors

## Acknowledgement

This research was conducted using data from the REasons for Geographic and Racial Differences in Stroke (REGARDS) study. The authors thank the investigators, staff, and participants of REGARDS for their valuable contributions. A complete list of participating REGARDS investigators and institutions can be found at: https://www.uab.edu/soph/regardsstudy/.com. The contents do not represent the view of the U.S. Department of Veterans Affairs or the United States Government.

The authors thank the other investigators, the staff and the participants of the REGARDS study for their valuable contributions.

## Statement of Ethics

The REGARDS study was approved by the institutional review boards of all participating institutions. A full list of sites and ethics committees is available at: https://www.uab.edu/soph/regardsstudy/. All participants provided written informed consent.

## Data Availability Statement

The data in this study was obtained from the REGARDS study, a third-party source, where access restrictions apply. Datasets may be requested from the REGARDS Study team at https://www.uab.edu/soph/regardsstudy/

